# Less is more in language production: Shorter sentences contain more informative words

**DOI:** 10.1101/2022.06.02.22275938

**Authors:** Neguine Rezaii, Boyu Ren, Megan Quimby, Daisy Hochberg, Bradford Dickerson

## Abstract

Agrammatism is characterized by short sentences, the omission of function words, a higher ratio of heavy to light verbs, and a decreased use of verbs relative to nouns. Despite the observation of these phenomena more than two centuries ago, there has been no unifying theory to explain all features of agrammatism. Here, by first examining the language of patients with primary progressive aphasia, we show that the seemingly heterogeneous features of agrammatism can be explained by a process that selects lower frequency words over their higher frequency alternatives in the context of a limitation in sentence production, likely to increase the informational content of sentences. We further show that when healthy speakers are constrained to produce short sentences, features of agrammatism emerge in their language. Finally, we show that these findings instantiate a general property in healthy language production in which shorter sentences are constructed by selecting lower frequency words.

---

Agrammatism is a disorder of sentence production characterized by short utterances, the omission of function words, and a decreased use of verbs relative to nouns (1–8). At the severe stages of the disorder, the language output reduces to single word utterances that are mainly nouns (9). The agrammatic style of language production that is often found in patients with non-fluency has been classically described in patients with ischemic strokes or other lesions in Broca’s area and more recently in neurodegenerative diseases such as Primary Progressive Aphasia (PPA) (for a comparison see (10–15)). After more than two centuries since the observation of these phenomena (16) and despite the succession of accounts aiming to explain this phenomenon, there has yet been no single theory to account for all features of agrammatism. Instead, each feature has been explained through a distinct neurolinguistic process, largely independent of other features.

One of the most consistently reported features of agrammatism is the omission of function words whether in free forms such as pronouns, auxiliary verbs, conjunctions or bound forms such as inflectional changes to a word (4, 5, 17–19). In contrast to content words that carry the main message of a sentence (20), function words primarily play a grammatical role with likely distinct storage and access processes than those of content words (21, 22). As agrammatism disrupts the syntactic structure of a sentence, access to function words has been hypothesized to be more affected than access to content words resulting in a reduced function-to-content words ratio (23–25). Under this dual system proposal, however, it remains unclear why agrammatic patients tend to use verbs in gerundive forms, a verb form that, at least in English, requires additional inflectional changes (26, 27, 7, 28, 3).

The other key symptom of agrammatism is an increased use of nouns over verbs (29–37). As both nouns and verbs are content words, the dissociation in their use cannot be explained by the dual retrieval process that occurs for function and content words. Instead, it has been hypothesized that the verb deficit is related to the greater syntactic complexity of verbs relative to nouns as verbs implicitly process the syntactic relations among elements of a sentence (6, 28, 31, 38, 39). Relatedly, a neurobiologically-driven hypothesis postulates that a deficit in processing actions affects verbs more than nouns and that verb impairments are usually associated with anterior brain lesions, which lie close to motor cortex, while noun impairments are more often associated with posterior lesions, which lie close to sensory association areas (29, 40–43). Other researchers reject the syntactic processing disorder hypothesis, arguing that the dissociation in use of nouns and verbs in agrammatism is one of lexical processing involving the category of verbs based on a proposal in which the lexicon is organized along the lines of the distinct noun and verb classes (6, 32, 33, 44, 45).

A more recent case of a lexical dissociation in agrammatism is a decrease in use of light verbs relative to heavy verbs when compared with healthy language production (46, 47). Light verbs such as *go, make, do, get*, and *take* are semantically more general and associated with less specific objects (48–50). Breedin and colleagues (1998) hypothesized that heavy verbs are more resistant to disruption as they are semantically richer and more specific to a particular context than light verbs. A lexical dissociation for heavy/light verbs represents yet another case of lexical dissociation independent from that of content/function words and nouns/verbs (51).

In the absence of a theory explaining the features of agrammatism under a single account, the field has proceeded with the assumption that agrammatism is a multi-component deficit disorder in which a distinct mechanism in disrupted in each component of the deficit (6). In this context, the co-occurrence of the various agrammatic symptoms is likely due to the neural proximity of the processes involved in each symptom (6). In this work, we propose a view that the agrammatic style of language production is the outcome of selecting words of lower frequency over other alternatives at a given lexical decision point. Under this hypothesis, the tendency to use content words, nouns, and heavy verbs in an utterance is likely due to their lower word frequency when compared with their alternatives–respectively, function words, verbs, and light verbs. As the lower probability of occurrence of a word has an inverse relation with the information it contains (52), their use by agrammatic patients could potentially be viewed as a compensatory mechanism to increase the informational content of shorter sentences.

To test this hypothesis, we examine the language of patients with primary progressive aphasia (PPA), a neurodegenerative disorder that predominantly affects language (53, 54), which has three major variants. The non-fluent variant (nfvPPA) is characterized by agrammatic and/or nonfluent speech, the logopenic variant (lvPPA) by deficits in word retrieval and sentence repetition, and the semantic variant (svPPA) by impairments in single word comprehension and confrontational naming (55). We first evaluate the language samples of patients with nfvPPA to show that in each case of lexical dissociation described above, the selected lexical item is of lower frequency than its alternative word type. We then show that this potential compensatory strategy in agrammatism originates from a basic property in normal language production by asking healthy speakers to produce short utterances of one to two words. Under this constrained condition, we expect the language of healthy speakers to show features similar to those of patients with agrammatism. Next, we test the hypothesis that as a basic property of language production in healthy speakers, shorter sentences contain words with lower frequency (often more specific informational content). Finally, we evaluate the language of patients with lvPPA and svPPA to see whether the relationship between sentence length and word frequency is disrupted in patients who have difficulty accessing low frequency words (56– 58).

## Methods

### Participants

One hundred and one patients with PPA were recruited from an ongoing longitudinal study being conducted in the Primary Progressive Aphasia Program in the Frontotemporal Disorders Unit of Massachusetts General Hospital (MGH). Baseline clinical and language assessments were used to characterize and subtype patients into nfvPPA (n = 35), svPPA (n = 25), and lvPPA (n = 41). The participants of this study underwent a comprehensive clinical evaluation as previously described (59). The evaluation included a structured interview by a behavioral neurologist or neuropsychiatrist and a neurological examination as well as speech and language assessment by a speech-language pathologist. The protocol for the participants of this study included the National Alzheimer’s Coordinating Center (NACC) Uniform Data Set measures. We also include ratings on our scale called the Progressive Aphasia Severity Scale (PASS). Modeled after the Clinical Dementia Rating Scale, the PASS uses the clinician’s best judgment, integrating information from the patient’s examination and test performance in the office as well as a companion’s description of routine daily functioning (60). The PASS includes “boxes” for fluency, syntax, word retrieval and expression, repetition, auditory comprehension, single word comprehension, reading, writing, and functional communication. The PASS, Sum-of-Boxes is a summary measure of aphasia severity (a sum of each of the box scores). The clinical and demographic information on the patients is shown in table 1. Thirty-three age matched healthy controls were included in the first part of this study enrolled through the Speech and Feeding Disorders Laboratory at the MGH Institute of Health Professions. These participants passed a cognitive screen, were native English speakers, and had no history of neurologic or developmental speech or language disorders. All clinic participants gave written informed consent in accordance with guidelines established by the Mass General Brigham Healthcare System Institutional Review Boards which govern human subjects research at MGH. This study was approved by the BRAINS at MGH.

**Table 1.** Clinical and demographic characteristics of participants performing the unconstrained task

|  | nfvPPA | lvPPA | svPPA | controls |
| --- | --- | --- | --- | --- |
| Sample size | 35 | 41 | 25 | 33 |
| Mean age (SD) | 70.3 (9.3) | 70.7 (6.2) | 66.2 (7.6) | 64.9 |
| Handedness, right | 89% | 84% | 88% | 74% |
| Mean years of education (SD) | 17.3 (7.4) | 16.6 (2.2) | 16.6 (1.7) | 15.8 |
| Gender, female | 60% | 44% | 60% | 56% |
| PASS Sum-of-Boxes | 5.7 (3.6) | 5.8 (2.6) | 5 (2.2) | - |
| SD: standard deviation, PASS: Progressive Aphasia Severity Scale |  |  |  |  |

For the constrained language task, a separate cohort of 31 individuals were recruited from Amazon’s Mechanical Turk (MTurk). MTurk participants filled out a short survey about their neurological and language background. Only language samples from participants who were native English speakers, with no self-reported history of brain or speech-language disorder, either developmental or acquired, were included in the analyses. These individuals had an average age of 47.6 with an average year of education of 16.1. 71% of them were female and 88% were right-handed.

### Language samples

The participants were asked to look at a drawing of a family at a picnic from the Western Aphasia Battery– Revised (61) and describe it using as many full sentences as they could. Responses were audio-recorded using an Olympus VN-702PC Voice Recorder (Center Valley, PA, USA) in a quiet room and later transcribed into text using Microsoft Dictate application. The transcriptions were then manually checked for accuracy by a research collaborator who was blind to the grouping. Disfluencies of speech such as repetitions and use of fillers, such as “um”, “you know”, etc., were identified per the protocol previously described (62) and removed from further analyses.

For the constrained condition, the Mturk participants were asked to describe the same picture using either one word or two-word sentences only.

As language data was sparse for sentences containing more than 20 words (about 1% of all utterances), we report the results based on about 99% of the data (2289 of the total of 2314 sentences) with sentence length of less than and equal to 20.

### Text analysis of language samples

We used Quantitext, a text analysis toolbox we developed in Frontotemporal Disorders Unit of Massachusetts General Hospital, to automatically produce a set of quantitative language metrics. The goal of developing this package is to increase the precision and objectivity of language assessments while reducing human labor (as outlined in (63)). The toolbox uses a number of natural language processing toolkits and software such as Stanford Parser (64), spaCy (65), and NLTK as well as text analysis libraries in R. Quantitext receives transcribed language samples as input and generates as outputs a number of metrics such as sentence length, log word frequency, log syntax frequency (63), content units (as in (66)), efficiency of lexical and syntactic items (as in (67)), part of speech tags, and the distinction of heavy and light verbs.

### Measuring word frequency

To measure word frequency, we used the Switchboard corpus (68), which consists of spontaneous telephone conversations averaging 6 minutes in length spoken by over 500 speakers of both sexes from a variety of dialects of American English. We use this corpus to estimate word frequency in spoken English, independently of the patient and control sample. The corpus contains 2,345,269 words. Our analyses consider sentences, not words, as a basic unit. The word frequency of each utterance was calculated by taking the average log frequency of all words within a sentence based on the Switchboard corpus.

### Content to all word ratio

The part of speech of each word was automatically determined by Quantitext. Nouns, verbs (except *be, have* and *do*), adjectives and adverbs were considered as content words. All other words were classified as function words. Here, we measured content to all word ratio by dividing the number of content words by the number of all words in a sentence.

### Noun to verb ratio

The noun to verb ratio was measured by dividing the number of nouns by the sum of nouns and verbs in each sentence.

### Heavy verb to all verb ratio

The following verbs were classified as light verbs: ‘go’, ‘have’, ‘do’, ‘come’, ‘give’, ‘get’, ‘make’, ‘take’, ‘be’, ‘bring’, ‘put’, ‘move’ while excluding auxiliaries from this list (47). All other verbs were classified as heavy verbs. Heavy verb to all verb ratio was measured by dividing the number of heavy verbs by the total number verbs in a sentence.

### Statistics

For the statistical analyses of this study, we used the R software version 4.1.2. To compare the features of agrammatism across different groups, we used mixed-effects models with subject-specific random intercept via the lme4 package in R (69). We used independent t-tests to compare the occurrences of lexical items belonging to different parts of speech in the Switchboard corpus.

To estimate the smooth but potentially nonlinear relationship between sentence length and word frequency, we used generalized additive models (GAM). GAM is a type of generalized linear model in which the mean of the outcome is a sum of unknown smooth univariate functions of continuous predictors (70). These unknown smooth functions can be estimated either parametrically (e.g., via a series of basis functions) or nonparametrically (e.g., kernel smoothing) (71). Spline functions are popular choices for bases in GAM due to their ability to approximate any smooth function when the number of internal knots is large enough (72). To avoid overfitting and promote generalizability of the fitted model, a smoothness penalty on the spline function is usually employed to prevent the model from interpolating. A commonly used class of penalties targets the L2 norm of the derivative of a given order and controls the complexity of the fitted GAM. We use thin plate regression splines (73) as the basis functions and set the number of internal knots of the spline to be adequately large (e.g. 50). The value of effective degrees of freedom (EDF) formed by the GAM model shows the degree of curvature of the relationship. A value of 1 for EDF is translated as a linear relationship. Values larger than one denote a more complex relationship between the predicting and outcome variables. We used the “gam” function in the “mgcv” package in R to fit the model (74). We included in the model separate spline functions of sentence length for each group of subjects (e.g., PPA variants vs. healthy controls) and a subject-specific random slope. The model parameters were estimated via restricted maximum likelihood (REML) method (75). To test whether the relationship between word frequency and sentence length were different in PPA variants when compared to healthy controls, we performed a generalized likelihood ratio test for penalized splines (76). An alpha value of 0.05 was set as our a priori threshold for statistical significance, with values below 0.1 indicating a trend-level effect.

## Results

### 1. Patients with nfvPPA exhibit agrammatic speech production and produce lower frequency words than controls

1.1. Here, we first establish that our cohort of patients with nfvPPA exhibit the canonical features of agrammatism. We fit a mixed-effects model with subject-specific random intercept to predict each feature of agrammatism (sentence length, function to all word ratio, noun to verb ratio, heavy to all verb ratio, and verbs in gerund form to all verb ratio) at the sentence level with group as a predictor. Compared to healthy controls, patients with nfvPPA used shorter sentences as measured by the number of words per sentence (β=-2.975, SE= 0.454, t=-6.548, p<0.001) (see Supplementary Table S1).

Regarding the lexical dissociations in agrammatism, nfvPPA patients used a higher content word to all word ratio (β = 0.089, SE= 0.027, t= 3.295, p = 0.002), a higher noun to verb ratio (β=0.063, SE= 0.025, t=2.534, p=0.014), and a higher heavy verb to all verb ratio (β=0.175, SE= 0.044, t=3.952, p<0.001) when compared with healthy controls. As will be shown in section 1.2, in all three of these cases, nfvPPA patients tend to produce a word class that has a lower frequency in healthy people’s speech than its alternative word class. In addition, the language of nfvPPA patients showed a trend toward higher gerund to all verb format ratio (β=0.076, SE= 0.044, t=1.729, p=0.089) than healthy controls. The mean and standard deviation of the features of agrammatism in healthy controls and nfvPPA patients are provided in Table S1 of the Supplementary Material. Figure 1 compares the features of agrammatism at the subject level between healthy controls and patients with nfvPPA.

**Figure 1.**
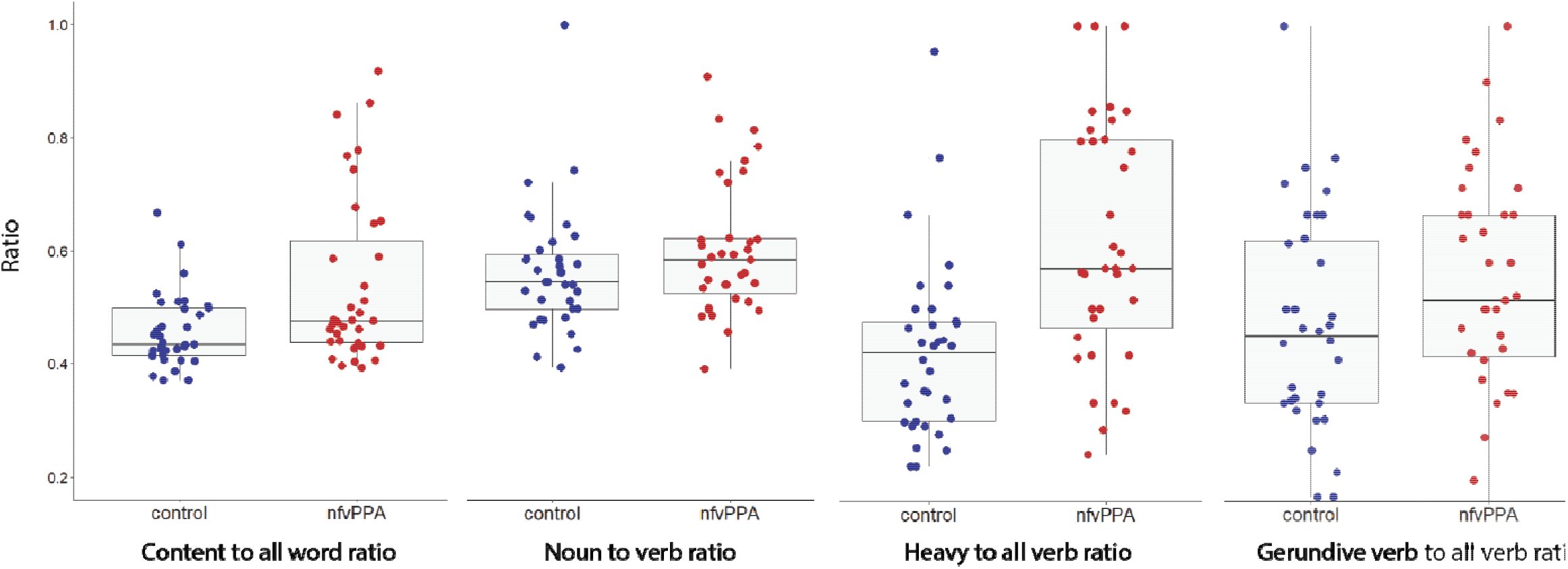
Box plots of features of agrammatism (content to all word ratio, noun to verb ratio and heavy to all verb ratio, gerundive verb to all verb ratio) at the subject level in healthy controls and nfvPPA patients. The boxes show 25^th^, 50^th^, and 75^th^ percentile and the whiskers represent the minimum and maximum values, excluding outliers.

1.2. Next, we analyzed the Switchboard corpus to compare the occurrence of function words with content words, nouns with verbs, and heavy verbs with light verbs in the everyday language of English speakers. Using independent t-tests, we found that content words appear less frequently than function words (t(34,656) = -5.5825 p<0.001), nouns less than verbs (t(25,453) = -3.9716, p < 0.001), and heavy verbs less than light verbs (t(5719) = -3.0149, p < 0.001). The mean and standard deviation of occurrences of each word class is provided in Table S2 of the Supplementary Material.

1.3. Based on these two lines of evidence, we tested the hypothesis that patients with agrammatism use words with a lower frequency than healthy controls. Fitting a mixed-effects model with random effects for subjects to predict word frequency with subject group (treatment-coded with healthy controls as reference level) as a predictor, we found that the average word frequency of a sentence produced by nfvPPA patients (mean = 7.4, SD = 1.7) is lower than that of controls (mean = 8.2, SD = 0.8) (β=-0.617, SE= 0.158, t=-3.897, p<0.001).

### 2. In sentences from healthy individuals constrained to produce short sentences, features of agrammatism emerge

Here, we compare the constrained and unconstrained language production of healthy speakers with respect to the features of agrammatism. Fitting a mixed-effects model with random effects for subjects with language production condition as a predictor (treatment-coded with unconstrained production as reference level), we found that constrained language production in healthy individuals contained a higher content to all word ratio (β = 0.534, SE = 0.011, t=-49.96, p<0.001), a higher noun to verb ratio (β=0.091, SE= 0.028, t=3.295, p=0.001), a higher heavy verb to all verb ratio (β=0.565, SE= 0.032, t=17.47, p<0.001), and a higher gerund to all verb format ratio (β=0.117, SE= 0.051, t=2.288, p=0.024). Constrained language also contained lower frequency words (β=-3.439, SE= 0.090, t=-38.17, p<0.001) and lower frequency content words (β=-0.705, SE= 0.116, t=-6.075, p<0.001). The mean and standard deviation of the features of agrammatism under the unconstrained and constrained conditions are provided in Table S3 of the Supplementary Material. Figure 2 compares the features of agrammatism at the subject level between the unconstrained and constrained samples in these healthy individuals.

**Figure 2.**
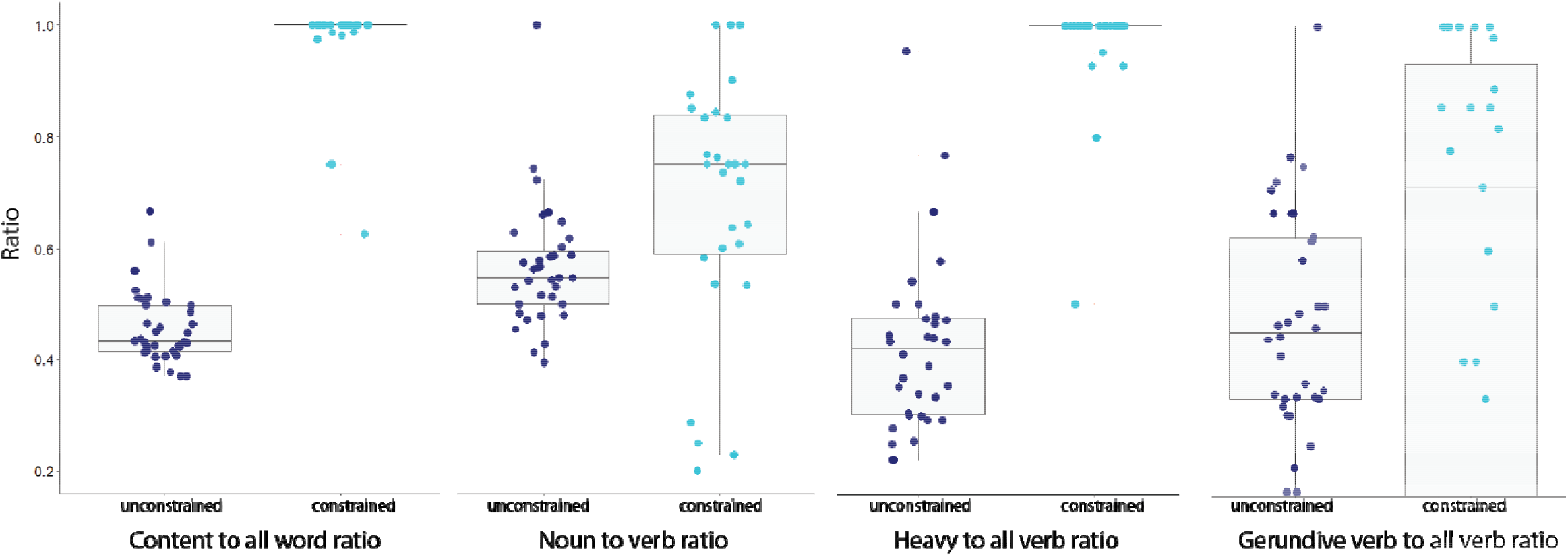
Box plots of features of agrammatism (content to all word ratio, noun to verb ratio and heavy to all verb ratio, gerundive verb to all verb ratio) at the subject level in the unconstrained and constrained language samples of healthy speakers. The boxes show 25^th^, 50^th^, and 75^th^ percentile and the whiskers represent the minimum and maximum values, excluding outliers.

**Figure 3.**
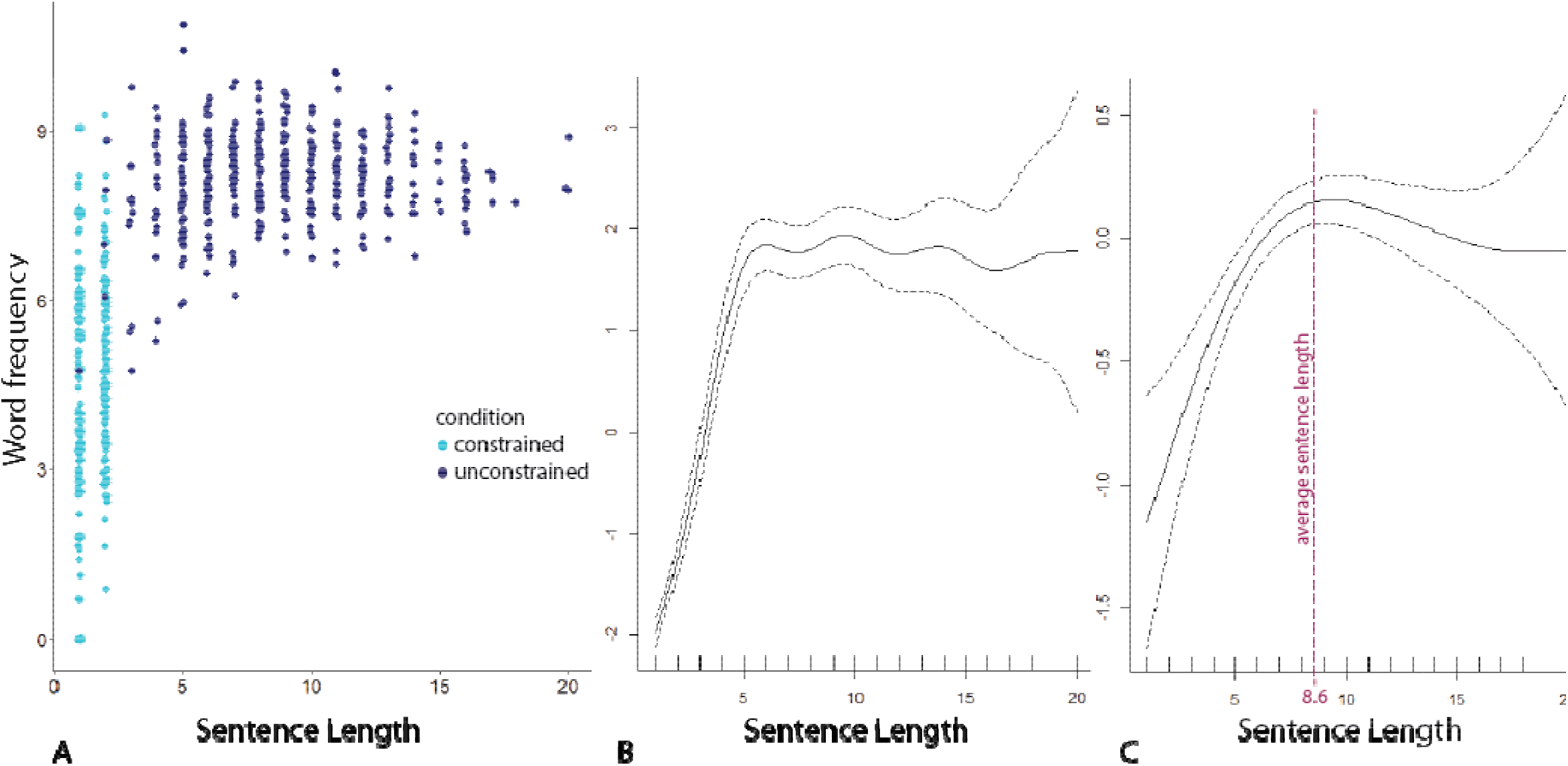
**A**. Scatterplot showing the relationship between sentence length and word frequency under unconstrained and constrained conditions in healthy individuals. **B**. GAM plot showing the relationship between sentence length and word frequency under unconstrained and constrained conditions in healthy individuals. **C**. GAM plot showing the relationship between sentence length and word frequency of unconstrained language production in healthy individuals. The vertical line indicates the average sentence length in unconstrained language production. The two dashed lines in panel **B** and **C** indicate the pointwise 95% confidence intervals of the fitted curves.

### 3. Shorter sentences generally contain lower frequency words

#### 3.1. Shorter sentences generally contain lower frequency words in the sentences of healthy speakers

We then tested the general hypothesis that shorter sentences contain words with lower frequency in healthy speakers. To cover a broad range of sentence lengths (including one- and two-word sentences), we combined the constrained and unconstrained results in healthy controls as shown in Figure 2A. We fitted a GAM to this combined dataset with a subject-specific random intercept to model the relationship between sentence length and word frequency. We found that the average frequency of all words could be predicted from the length of a sentence (EDF = 7.2, p<0.001) as shown in Figure 2B. We also fitted a GAM with random intercepts to the data for unconstrained language production. A similar pattern was seen in the unconstrained language production of the healthy controls in which the average frequency of all words could be predicted from sentence length (EDF = 3.55, p<0.001) as shown in Figure 2C.

As shown in figures 2B and 2C, the sentence length-word frequency relationship is approximately linear until word frequency starts to plateau. To determine the sentence length at which the plateau starts to occur, we created a random data set where the value of sentence length varied from 1 to 20 with 0.1 increments. We then used the fitted GAM of sentence length-word frequency in the unconstrained healthy language production data to predict the word frequency of the randomly created data set. We found that the maximum word frequency occurs at a sentence length of 9.4.

Importantly, the average sentence length of unconstrained healthy language production is 8.6, suggesting that word frequency plateaus at about the average sentence length, the vertical dashed line in Figure 2C.

#### 3.2. Shorter sentences generally contain lower frequency words in the utterances of all three variants of PPA, but with a bias toward higher frequency words in lvPPA and svPPA

Here, we evaluate the word frequency and sentence length relationship in the three variants of PPA. In each of the three variants, the average frequency of all words could be predicted from sentence length (all p-values < 0.001).

To test whether the relationships between word frequency and sentence length are different in PPA variants when compared to healthy controls, we performed a generalized likelihood ratio test for penalized splines and examined whether there was an interaction between sentence length and group. We added each of the PPA variants to the data from healthy speakers one at a time to compare the full model that contained variant as a predictor with the null model. We found that the sentence length-word frequency relationship in healthy speakers was different from that of lvPPA and svPPA (Df = 21.0, Deviance = 156.1, p < 0.001 and Df = 151.4, Deviance = 171.3, p < 0.001 respectively), but not different from that of nfvPPA (Df = 46.7, Deviance = 45.6, p = 0.999).

As shown in Figure 4A, lvPPA and svPPA show a similar sentence length-word frequency relationship to healthy controls but with an upward shift (a bias toward higher frequency words). That is, although the use of shorter sentences predicts lower frequency words in each group, lvPPA and svPPA produce relatively higher frequency words at any given sentence length.

**Figure 4.**
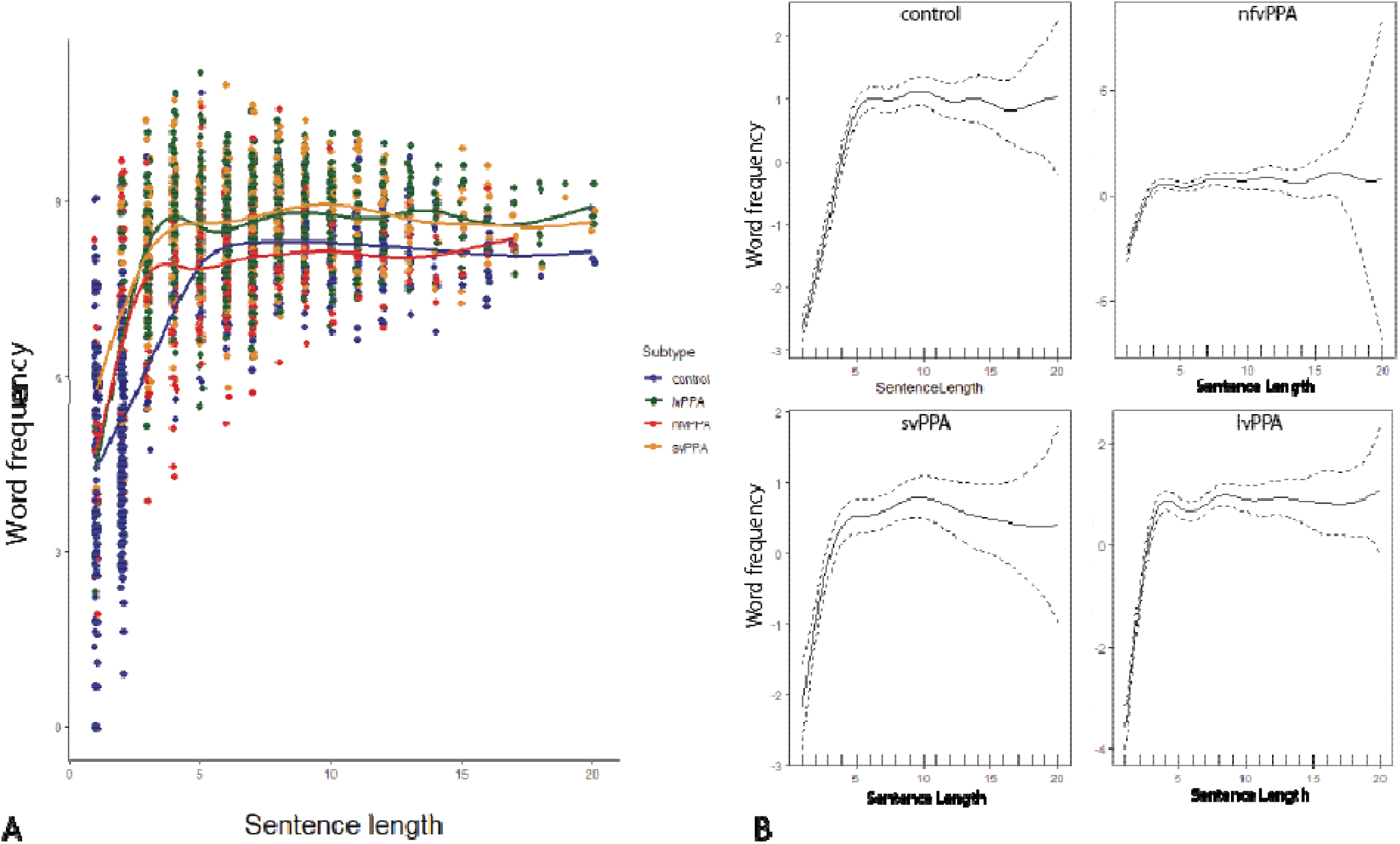
**A**. Scatterplot showing the relationship between sentence length and the average frequency of all words at the sentence level in all four groups. **B**. GAM plots of all smooth terms where the two dashed lines indicate the pointwise 95% confidence intervals of the fitted curves.

#### 3.3. Shorter sentences contain lower frequency content words in all three variants of PPA, but with a bias toward higher frequency words for lvPPA and svPPA

Here, we test whether the average frequency of content words (as opposed to all words in a sentence) holds a similar relationship with sentence length. Fitting a GAM on all data from healthy controls, both constrained and unconstrained to cover a wide range of sentence lengths (with a random intercept for subject) showed a significant relationship between content word frequency and sentence length (p < 0.001). As shown in Figure 5, there is a positive relationship between sentence length and content word frequency up to a point where the data begin to plateau.

**Figure 5.**
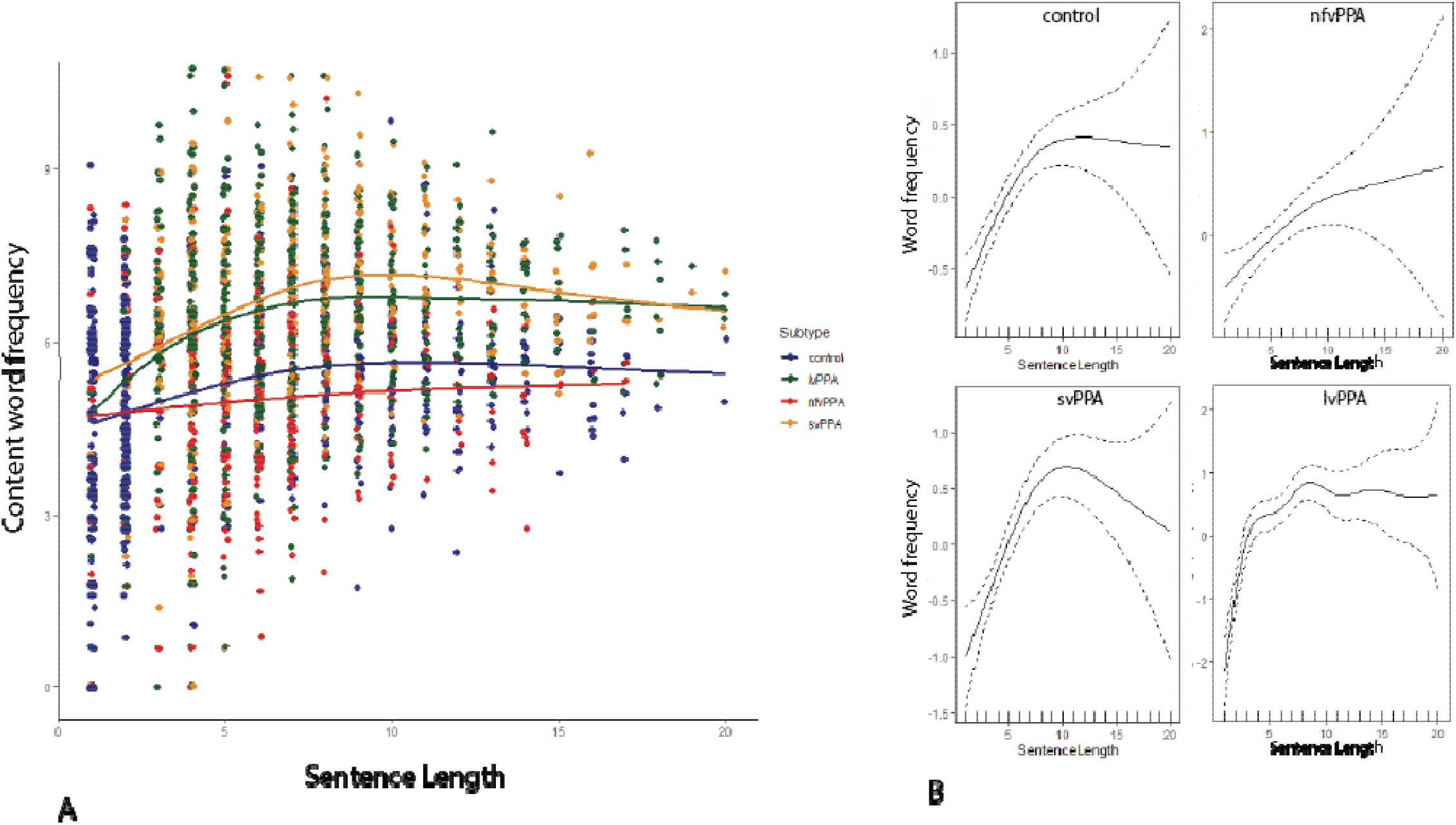
**A**. Scatterplot showing the relationship between sentence length and the average frequency of content words at the sentence level. **B**. GAM plots of all smooth terms where the two dashed lines indicate the pointwise 95% confidence intervals of the fitted curves.

We used a similar test as in 3.2 to examine whether the relationship between content word frequency and sentence length are different in PPA variants when compared to healthy controls. The results suggest that the sentence length-content word frequency relationship in healthy speakers was different from that of lvPPA and svPPA (Df = -7.2, Deviance = -75.3, p < 0.001 and Df = -0.1, Deviance = -4.6, p = 0.02 respectively), but not different from that of nfvPPA (Df = 4.0, Deviance = 21.2, p = 0.08).

#### 3.4. Controlling for function to all word ratio in the sentence length-word frequency relationship

As longer sentences tend to contain more function words, we evaluated the sentence length-word frequency relationship by further controlling for the function to all word ratio (i.e. by adding function to all word ratio to the statistical model). Here, we use a multivariable GAM to predict the average frequency of all words of a sentence from its length and function word to all word ratio with a subject-specific random intercept in a dataset that combined all groups. We found a significant relationship between sentence length and the average frequency of all words (EDF = 6.4, p < 0.001) as well as function word to all word ratio (EDF = 2.5, p < 0.001). Figure 6 indicates that the average frequency of words in a sentence increases as sentence length and function to all word ratio increase.

**Figure 6.**
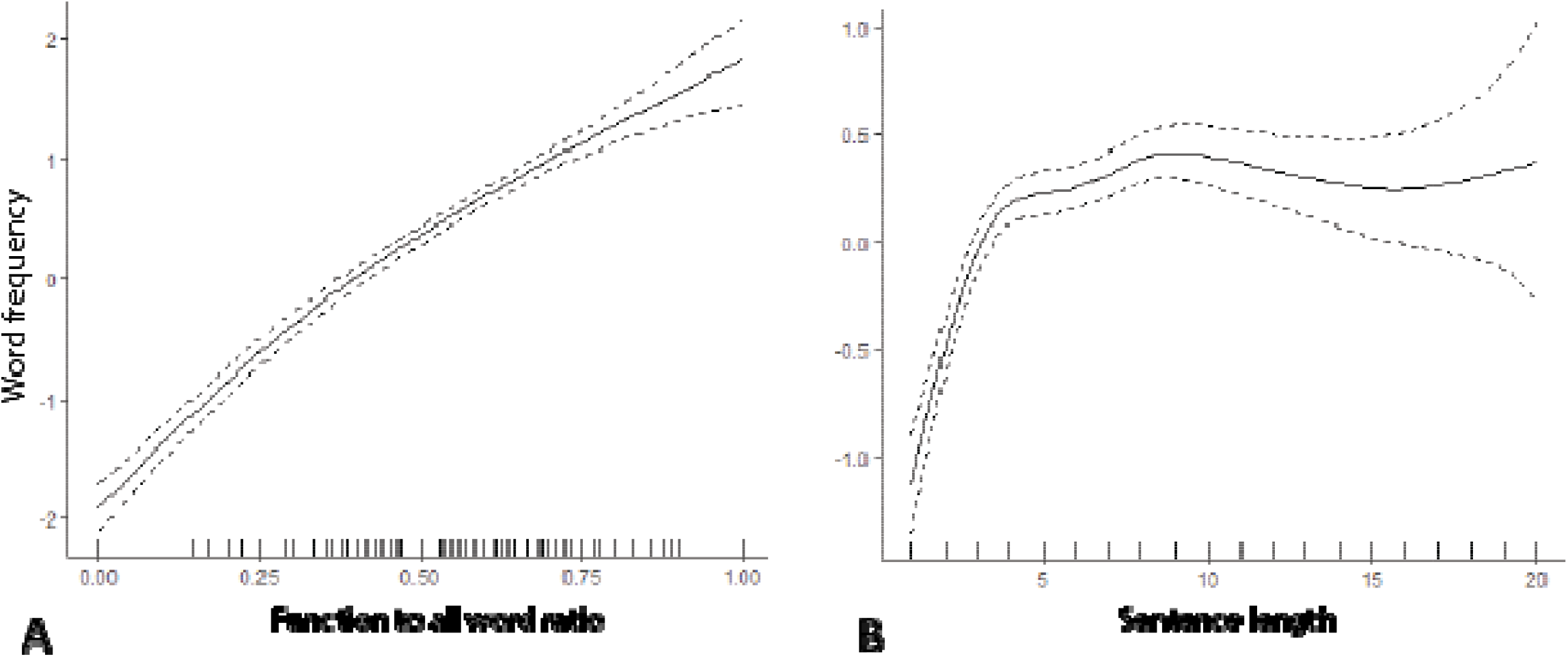
GAM plots of the two smooth terms in the model that predicts the average frequency of all words from **A**. function to all word ratio and **B**. sentence length.

## Discussion

We conducted this study to test our hypothesis that the seemingly heterogeneous features of agrammatism can be explained by a process in which lower frequency words are selected over their higher frequency alternatives in the context of a limitation in sentence production. The results of the study in aphasic patients support this hypothesis. Furthermore, the experiments in healthy individuals suggest that this is a fundamental property of normal language, albeit without the fixed limitation in sentence production that agrammatic aphasic patients experience. From the perspective of an information theoretic approach, the findings in agrammatic aphasic patients could potentially be interpreted as a compensatory mechanism in which more informative words, as measured by their lower probability of occurrence, when aphasia limits sentence production to shorter sentences (52).

In line with the idea that the features of agrammatism are the consequences of a compensatory mechanism, Kolk (1985) had previously put forward the adaptation theory for the case of omission of function words (77). According to this theory, function words are dropped not because of an impairment in their use but as an adaptation to a delay in the process of sentence production. According to this view, the time-sensitive process of sentence production requires all elements of a sentence to be retained in memory so that function words can be put in the right place and the right morphological changes can be made. If the process becomes too slow, the representation of sentence elements will disappear from memory before the syntactic operations are finished which would lead to aborted sentences. Therefore, as an adaptation mechanism in patients with nonfluent speech, sentences become shorter by dropping function words so that syntactic operations take less time, and the delay exerts less of a blocking effect. The idea of compensation in agrammatism has also been entertained in Arnold Pick’s proposal on the economy of effort (78). According to this proposal, agrammatic patients try to restrict the consequences of their motor speech difficulties by producing only the most important words in the sentence, an adaptation strategy which leads to the omission of function words. Our work builds on these proposals and expands the idea of compensation to other types of lexical dissociations in agrammatism beyond the function/content word dissociation and operationalizes the measurement of the *important* (i.e., most specifically meaningful) words of a sentence.

If the features of agrammatism are interpreted as the outcome of a compensatory mechanism as proposed in this work, then a deficit in syntax processing would not be the central cause of the observed features. In this context, the term agrammatism could be considered a misnomer. This view is consistent with studies showing that the language of patients with agrammatism may follow the subject-verb-object construction (3). The absence of a central deficit in syntax processing also explains relatively intact verb comprehension as opposed to verb production (79–81), intact linguistic representation of and on-line access to the verb lexicon (82, 83), near normal performance on a grammaticality judgment task involving verb-argument-structure violations (79), and access to all possible argument structures of verbs in the immediate temporal vicinity of the verb during on-line sentence processing (82, 83). In addition, it has been shown that patients with agrammatism could still successfully recognize function words with normal reaction time despite dropping function words in their language production (77).

Our study further showed that this potential compensatory mechanism in agrammatism stems from a basic property of normal language production. When healthy speakers were faced with the constraint of producing only one- and two-word sentences, their language output exhibited features similar to those of patients with agrammatism, i.e., the ratio of content to all words, nouns to verbs, and heavy verbs to light verbs of their sentences would increase. Furthermore, healthy speakers used most verbs in gerundive form when restricted to produce one-to-two-word sentences. The increased use of gerunds is a well-known observation in agrammatism (26, 84–86). This runs counter to the observation that these patients have difficulty producing bound morphemes, since gerunds in some languages, including English, require inflectional changes. It has been proposed that the use of gerundive verbs is a shift to nominalize verbs to improve the access to verbs they are unable to produce (7, 28, 87) suggesting that their deficit does not affect the conceptual content of verbs but the grammatical category of verbs. The fact that under the constraint of producing short sentences, healthy speakers would also produce verbs mainly in gerund form suggests that the use of gerunds might be a succinct way of expressing the progressive aspect of a verb. For instance, in the picnic description task, both patients with agrammatism and healthy individuals under sentence length constraints use “boating” when they described the people on a sailboat in a lake.

Following the observation of selecting low frequency words in short sentences, we further tested the general hypothesis that the average word frequency of a sentence changes as a function of the sentence length. That is, up to a sentence length of about 9 words, shorter sentences contain more informative words. Crucially, as the average sentence length in the unconstrained speech of healthy speakers is also about 9 words, this sentence length-word frequency relationship may reflect the central tendency of an efficient tradeoff between the linguistic operations required to retrieve more specific words and to construct grammatically meaningful sentences.

The sentence length-word frequency relationship can be explained by the forces that shape human language to transfer the maximum amount of information with the least effort (88). In communicating a particular message, speakers have multiple options with regard to their choice of the words, syntax, and sentence length (89, 90). Yet many possible constructs would not be efficient—especially if they result in redundancy or verbosity. As a result, speakers tend to maintain a balance between the complexity of the various elements of their sentences, such as a balance between syntactic and lexical complexity for a uniform transfer of information (62). Speakers may also optimize the efficiency of communication by balancing the length of a sentence and the informational content of its words. Longer sentences require more cognitive effort to pronounce, write, read or interpret (91), as words at a longer distance from each other need to be linked together (92–94). As a result, speakers tend to choose the shortest possible sentence length from a potential set of sentences of approximately the same content (92, 95). On the other hand, the use of low frequency words requires greater cognitive effort as measured by longer reaction times (90, 96–98) or the extent of activated brain regions (99–105). As a result, if a neurologic condition imposes a limitation on the production of longer sentences, more cognitive resources would be available to allocate to retrieve low frequency words to convey the intended message.

Lastly, we showed that a similar relationship between sentence length and word frequency exists in the three variants of PPA. Patients with svPPA and lvPPA patients who are known to have difficulty accessing low frequency words continue to show the same relationship between sentence length and word frequency although with a bias indicating that the same sentence length-word frequency relationship is established at higher word frequencies. Since patients with nfvPPA do not have as much lexical retrieval difficulty for content words, their curve shows largely the same relationship between sentence length and word frequency as controls, although they choose slightly lower frequency words than controls at any sentence length. Future work is needed to determine the primary locus of the deficit such as poor executive function (10), impaired working memory, or deficient phonological processing (33, 106, 107) as the core mechanism underlying the fundamental limitation on sentence production in patients with “agrammatism.”

## Data Availability

All data produced in the present study are available upon reasonable request to the authors

## Acknowledgements

This research was supported by NIH grants R01 DC014296, R01 DC013547, R21 DC019567, R21 AG073744, and by the Tommy Rickles Chair in Primary Progressive Aphasia Research. We thank Dr. Jordan Green for sharing speech samples from control participants. We would like to express particular appreciation to the participants in this study and their family members, without whom this research would not have been possible.

