## Supplemental tables 1-3 for "Less is more in language production: Shorter sentences contain more informative words"

### Supplementary material

Table S1. Comparing features of agrammatism between healthy controls and nfvPPA patients

| Agrammatic features, Mean (SD) | Healthy controls | nfvPPA |
|---|---|---|
| Sentence Length | 8.60 (3.50) | 5.38 (3.23) |
| Content to all word ratio | 0.44 (0.14) | 0.54 (0.23) |
| Noun to verb ratio | 0.53 (0.21) | 0.61 (0.25) |
| Heavy to all verb ratio | 0.41 (0.35) | 0.56 (0.39) |
| Gerundive to all verb ratio | 0.41 (0.44) | 0.50 (0.46) |

Table S2. Mean and standard deviation (SD) of the occurrences (i.e. the counts) of each word class in Switchboard corpus

| Word class | Mean occurrence (SD) |
|---|---|
| Function | 538.24 (3841.53) |
| Content | 40.90 (611.77) |
| Noun | 21.93 (185.40) |
| Verb | 1253.00 (87.93) |
| Heavy verb | 59.69 (855.46) |
| Light verb | 13523.58 (15469.92) |

Table S3. Comparing features of agrammatism between unconstrained and constrained language production conditions in healthy speakers

| Agrammatic features, Mean (SD) | Healthy controls, unconstrained | Healthy controls, constrained |
|---|---|---|
| Sentence Length | 8.60 (3.50) | 1.41 (0.49) |
| Content to all word ratio | 0.44 (0.14) | 0.99 (0.07) |
| Noun to verb ratio | 0.53 (0.21) | 0.66 (0.36) |
| Heavy to all verb ratio | 0.41 (0.35) | 0.98 (0.15) |
| Gerundive to all verb ratio | 0.41 (0.44) | 0.70 (0.46) |
| Word frequency (log) | 8.2 (0.8) | 4.9 (1.9) |
| Content word frequency (log) | 5.5 (1.4) | 4.6 (1.9) |
